# Novel Oral Anticoagulants versus Vitamin K Antagonists for Patients with Left Ventricular Thrombus: A Systematic Review and Meta-Analysis

**DOI:** 10.1101/2020.12.08.20246330

**Authors:** Runzhen Chen, Jinying Zhou, Chen Liu, Peng Zhou, Jiannan Li, Ying Wang, Xiaoxiao Zhao, Yi Chen, Li Song, Hanjun Zhao, Hongbing Yan

## Abstract

**Background:** Although vitamin K antagonists (VKAs) are recommended as first-line anticoagulants for patients with left ventricular thrombus (LVT), accumulating evidence suggests novel oral anticoagulants (NOACs) could be safe alternatives for VKAs. Efficacy and safety of NOACs should be assessed to justify their usage for LVT patients.

**Design:** We performed a meta-analysis of observational studies to evaluate the efficacy and safety of NOACs as compared to VKAs in LVT patients.

**Methods:** PubMed and EMBASE databases were searched for articles published until November 12, 2020. Two reviewers independently extracted relevant information from articles and assessed the study quality. Pooled effects were estimated using Mantel–Haenssel method and presented as risk ratios (RR) using fixed-effect model. Reporting followed the Meta-analyses Of Observational Studies in Epidemiology (MOOSE) guideline.

**Results:** A total of 2467 LVT patients from 13 studies were included. Compared with VKAs, NOACs showed similar efficacy in prevention of stroke or systemic embolism (RR: 0.96, 95% confidence interval [CI]:0.80-1.16, P = 0.68) and thrombus resolution (RR: 0.88, 95% CI: 0.72-1.09, P = 0.20), but significantly reduced the risk of stroke (RR: 0.68, 95% CI: 0.47-1.00, P < 0.05). For safety outcomes, NOACs users showed similar risk of any bleedings (RR: 0.94, 95% CI: 0.67-1.31, P = 0.70), but lower risk of clinically relevant bleedings (RR: 0.35, 95% CI: 0.13-0.92, P = 0.03) compared with VKAs users.

**Conclusions:** Compared with VKAs, NOACs acquired similar efficacy and safety profile for patients with LVT, but could reduce the risk of strokes and clinically relevant bleedings.

---

Left ventricular thrombus (LVT) is a rare complication associated with acute myocardial infarction, heart failure and various cardiomyopathy ^1,2^. Due to the increased risk of embolic events, oral anticoagulation is required to prevent stroke or systemic embolism (SSE). Current guidelines recommend vitamin K antagonists (VKAs) for LVT patients based on limited evidence from observational studies ^1-4^. However, off-label use of novel oral anticoagulants (NOACs) for LVT is gaining interest, as they provide more constant anticoagulant effects and do not require continuous monitor of international normalized ratio (INR) ^1,5-9^. Numerous trials and analysis have already proved NOACs acquire similar efficacy in SSE prevention and lower bleeding risk in other clinical scenarios (e.g. atrial fibrillation, heart failure) as compared with VKAs ^10-12^, and there have been successful cases using NOACs to achieve complete LVT resolution and long-term favorable outcomes ^13,14^. Still, evidence is lacking for the use of NOACs in LVT patients. Therefore, this study aimed to summarize the up-to-date clinical findings comparing the efficacy and safety of NOACs and VKAs in LVT patients, in order to offer more insights for clinical practice and future randomized clinical trials (RCT) for anticoagulation of LVT.

## Methods

### Strategies for literature search

We performed a comprehensive literature search in Pubmed and EMBASE using the following search term: (“ventricular thrombi” or “ventricular thrombus”) and (“novel oral anticoagulants” or “direct oral anticoagulants” or “dabigatran” or “rivaroxaban” or “apixaban” or “edoxaban”) and (“vitamin k antagonists” or “warfarin” or “dicoumarol” or “phenindione” or “phenprocoumon” or “acenocoumarol” or “ethyl biscoumacetate” or “fluindione” or “clorindione” or “diphenadione” or “tioclomarol”). The literature search and data extraction process were completed by two researchers independently (R. Chen and J. Zhou), who are both physicians specializing in cardiology. The final search was completed on November 12, 2020.

### Study selection

Eligible studies must fulfill the following inclusion criteria: (a) patients were diagnosed with LVT by imaging evidence (e.g. transthoracic/transesophageal echocardiography, cardiovascular magnetic resonance imaging); (b) an RCT or observational study design; (c) comparing the outcomes of patients using NOACs vs VKAs. Studies regarding LVT secondary to ventricular device implantation, case reports, cases series, unpublished studies and studies not published in English were excluded in the current analysis. In case of missing data, authors of the original studies were contacted. A study would not be included in the pooled outcome analysis if the data for that specific outcome was unclear, not provided or failed to be acquired from contacting the original authors.

### Data extraction and quality assessment

The following data was extracted from the included study: surname of the first author, regions, study design, mean/median age or range of age, proportion of male sex, numbers of NOACs and VKAs users, numbers of outcomes, follow-up duration, primary causes of LVT and concomitant antiplatelet medications. Outcomes of interest included SSE, strokes, failures of thrombus resolution, any bleeding events and clinically relevant bleeding events (i.e. life-threatening bleedings, and bleedings requiring hospitalization or medical interventions). The quality of included studies was assessed by two independent authors (R. Chen and L. Song) using the Newcastle-Ottawa Scale (NOS). Should there be any discrepancies regarding data extraction and quality evaluation, a third author (J. Zhou) was consulted to reach a final consensus. Reporting followed the Meta-analyses Of Observational Studies in Epidemiology (MOOSE) guideline.

### Statistical analysis

RevMan 5.3 (The Cochrane Collaboration, Oxford, England) and Stata 15.0 (StataCorp, College Station, TX, USA) were used to perform this study. Pooled effects were estimated using Mantel– Haenssel method and presented as risk ratios (RR) with 95% confidence interval (CI). Heterogeneity across studies were examined using I^2^ statistics and chi-square Cochran Q test. An I^2^ > 50% or P < 0.1 for Cochran Q test signifies significant heterogeneity. A fixed-effect model was applied if no significant heterogeneity was observed; otherwise, a random-effect model was used. Funnel plots were used to detect potential publication bias. Begg’s rank correlation and Egger’s linear regression tests were performed when an outcome analysis included 10 or more studies. Trim-and-fill method was used to impute for missing studies and correct the publication bias with the *metatrim* command in Stata ^15^. A P-value < 0.05 was considered statistically significant.

## Results

### Studies characteristics and quality assessment

In the initial literature search, a total of 216 relevant records were identified after removing duplicates (**Figure 1**). After screening for titles and abstracts, 126 articles were excluded due to irrelevance. For the 90 articles entering eligibility assessment, 77 studies were further eliminated due to publication types or study design. Finally, thirteen articles were included in the synthetic analysis ^5-9,16-25^, two of which being prospective study. The average NOS scores was 6.2 (**Supplementary table 1**), showing that included studies were of median quality. A total of 2467 patients with LVT were included. The patients’ mean age ranged from 51.5 to 63.5 years, and the proportion of male patients was generally over 70%. The most common cause for LVT was ischemic heart diseases. The average follow-up duration ranged from 3 months to 3 years. Among users of NOACs, apixaban (50.0%) was most frequently prescribed, followed by rivaroxaban (40.8%), dabigatran (8.8%) and edoxaban (0.4%), while warfarin (98.4%) was predominantly prescribed to VKA users. Concomitant antiplatelet medication was prescribed in over half of the patients, but double antiplatelet treatment was less frequently used.

**Table 1.**
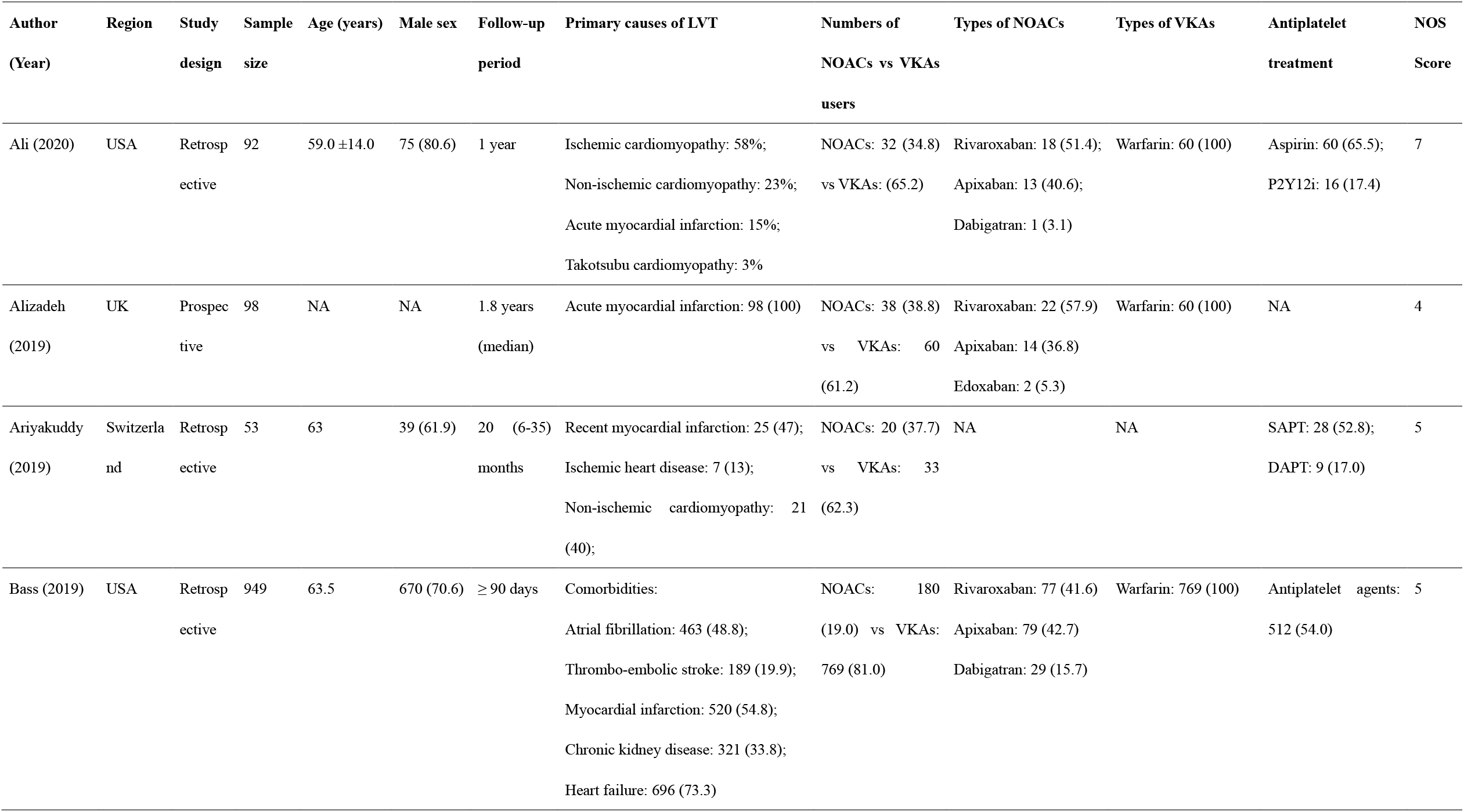
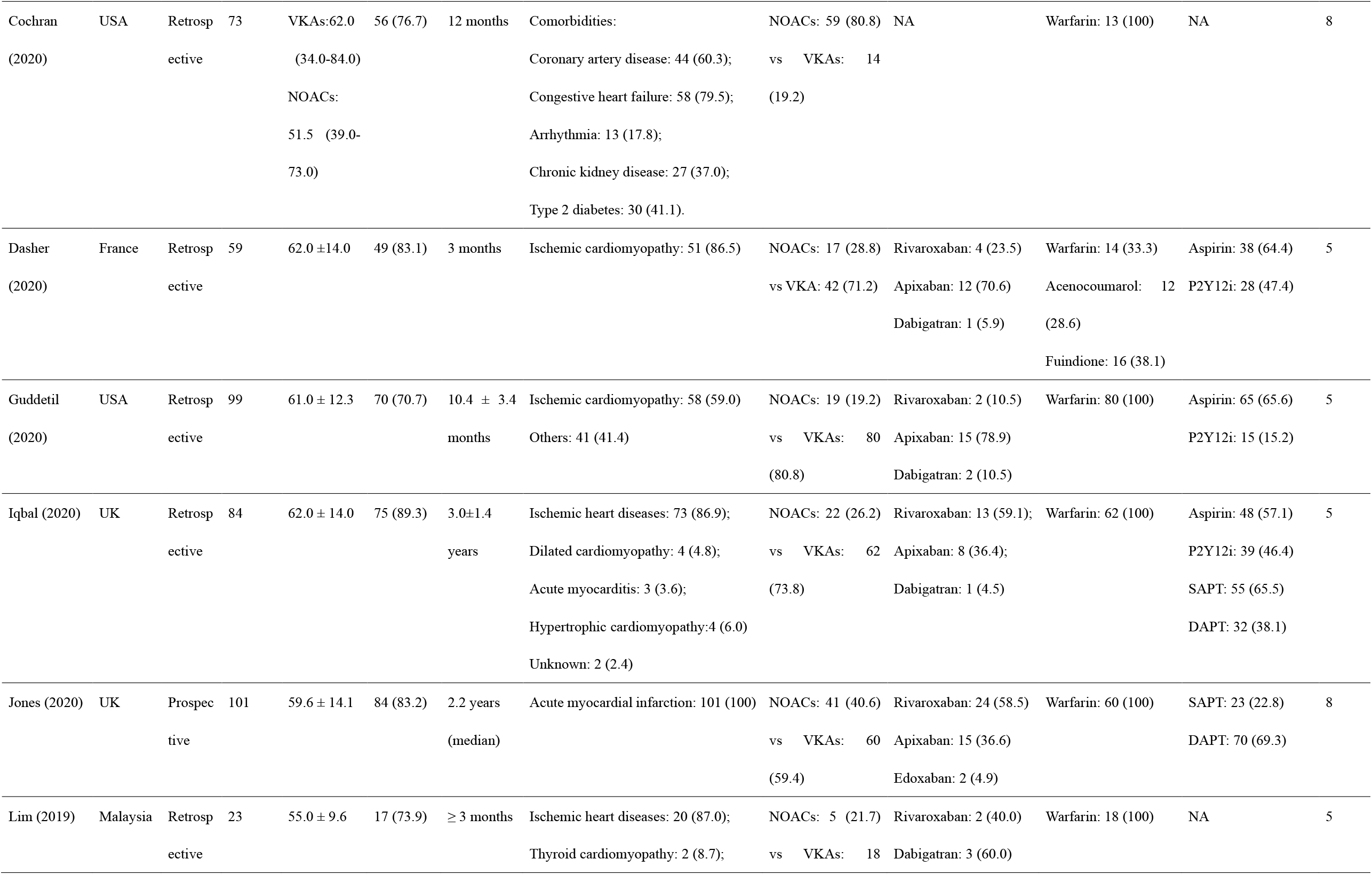
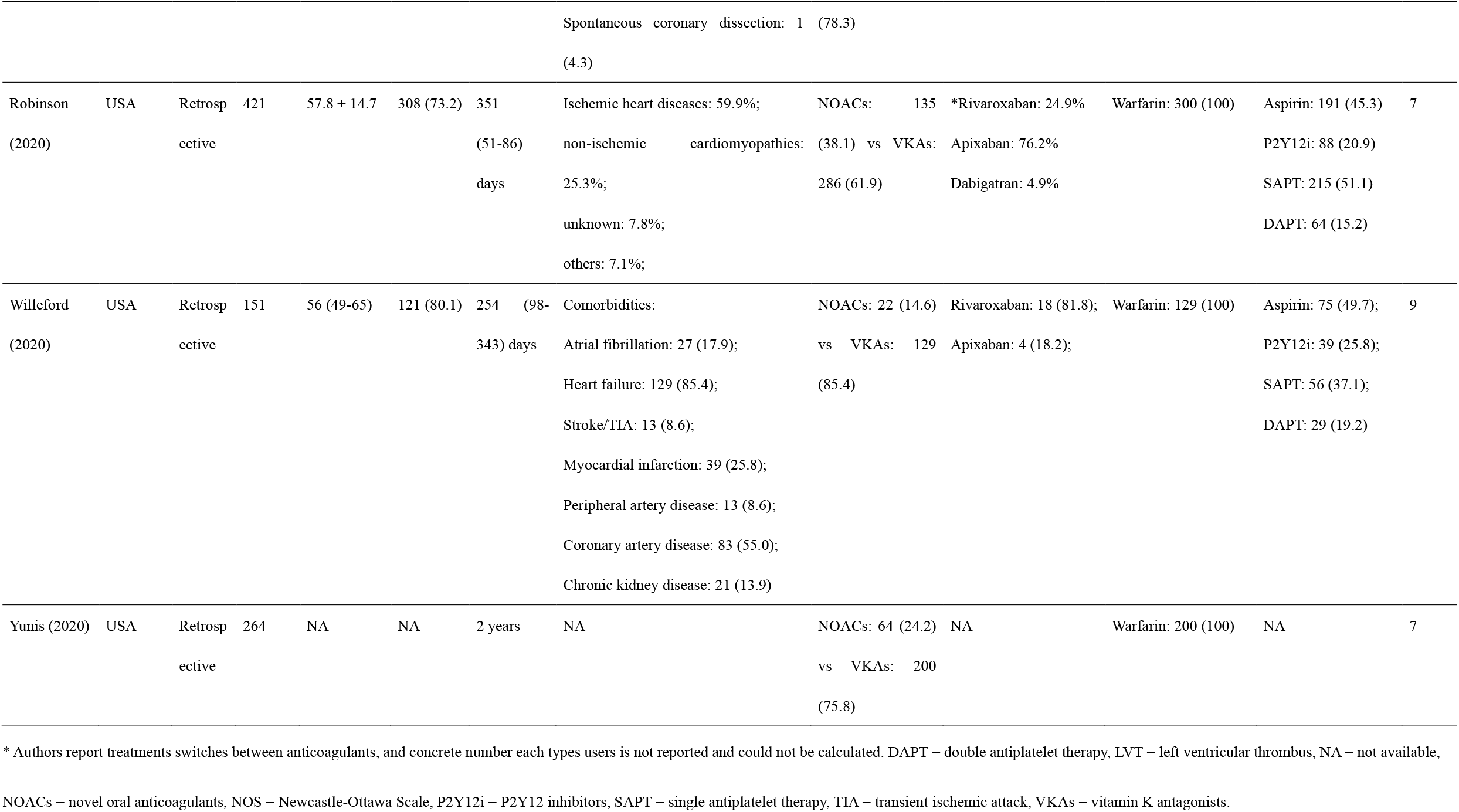
Essential characteristics of included studies.

**Figure 1.**
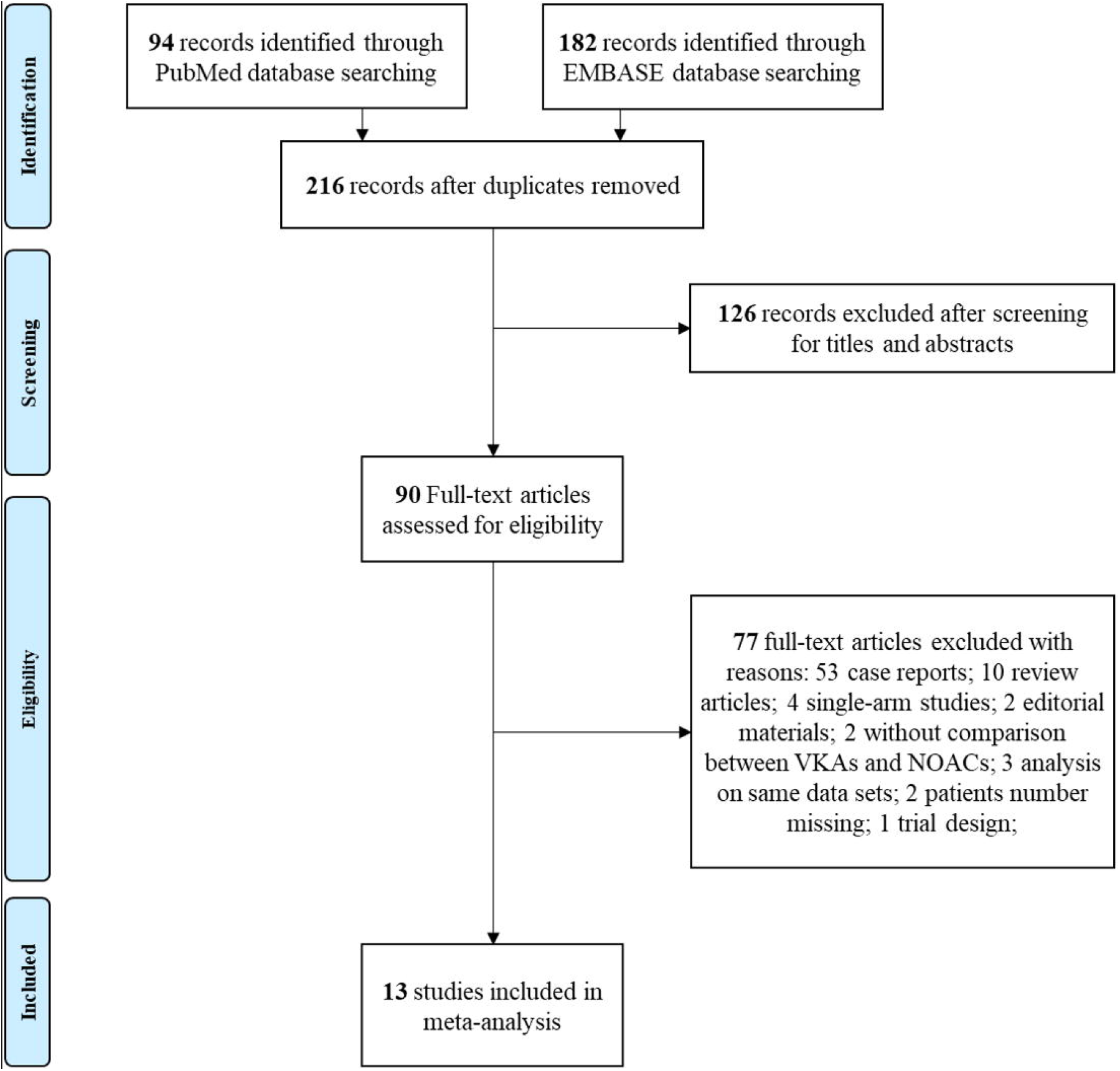
PRISMA flow diagram of study selection.

### Clinical outcomes

#### Stroke and embolic events

Ten studies reported the occurrence of SSE ^5-9,20-25^. The NOACs group and VKAs group did not show a significant difference in the risk of SSE (RR:0.96, 95% CI: 0.80-1.16, P = 0.69; I^2^ = 0%, **Figure 2A**). Publication bias was observed in funnel plots (**Supplementary figure 1A**) and statistical tests (Egger’s test, P = 0.029; Begg’s test, P = 0.733). After using trim-and-fill methods, the pooled effects remained similar (RR: 0.99, 95% CI: 0.83-1.20, P = 0.993), with no extra studies imputed (**Supplementary figure 2**). Notably, the study by Robinson et al reported treatment switches between NOACs and warfarin in 15.2% of patients (**Supplementary table 2**). Exclusion of this study led to similar results that NOACs acquired equivalent efficacy for SSE prevention as VKAs (RR: 0.95, 95% CI: 0.78-1.15, P = 0.59, I^2^ = 0). Meta-regression against age did not show significant impact on efficacy of NOACs and VKAs (RR: 1.06, 95% CI: 0.96-1.17, P = 0.225, **Supplementary figure 3A**). Subgroup analysis for follow-up duration (P _interaction_ = 0.07), sample size (P _interaction_ = 0.09), concomitant antiplatelet medication (P _interaction_ = 0.48) and primary causes of LVT (P _interaction_ = 0.09) showed a consistently similar effect of NOACs and VKAs (**Table 2**). Eight studies reported the outcome of strokes^6,9,19-22,24,25^. NOACs users showed lower risk of stroke compared with VKAs users (RR: 0.68, 95% CI: 0.68-1.00, P < 0.05, **Figure 2B**). Funnel plots showed no evidence of publication bias (**Supplementary figure 1B**). Meta-regression did not show significant impacts of age (RR: 1.01, 95% CI: 0.82-1.24, P = 0.899, **Supplementary figure 3B**). Subgroup analysis (**Table 2**) also showed consistent results across follow-up duration (P _interaction_ = 0.79), sample size (P _interaction_ = 0.49), concomitant usage of antiplatelet agents (P _interaction_ = 0.88) and primary etiologies for LVT (P _interaction_ = 0.76)

**Table 2.** Subgroup analysis of various outcomes for users of NOACs and VKAs.

| Subgroups | SSE |  |  |  |  | Stroke |  |  |  |  | Failure of thrombus resolution |  |  |  |  | Any bleedings |  |  |  |  | Clinically relevant bleedings |  |  |  |  |
| --- | --- | --- | --- | --- | --- | --- | --- | --- | --- | --- | --- | --- | --- | --- | --- | --- | --- | --- | --- | --- | --- | --- | --- | --- | --- |
|  | N | RR (95% CI) | I<sup>2</sup> | P | P<sub>int</sub> | N | RR (95% CI) | I<sup>2</sup> | P | P<sub>int</sub> | N | RR (95% CI) | I<sup>2</sup> | P | P<sub>int</sub> | N | RR (95% CI) | I<sup>2</sup> | P | P<sub>int</sub> | N | RR (95% CI) | I<sup>2</sup> | P | P<sub>int</sub> |
| All | 10 | 0.96 (0.80-1.16) | 0% | 0.68 | - | 8 | 0.68 (0.47-1.00) | 0% | 0.05 |  | 11 | 0.88 (0.72-1.09) | 26% | 0.2 | - | 9 | 0.94 (0.67-1.31) | 24% | 0.7 | - | 6 | 0.35 (0.13-0.92) | 0% | 0.03 | - |
| Follow-up duration |  |  |  |  |  |  |  |  |  |  |  |  |  |  |  |  |  |  |  |  |  |  |  |  |  |
| ≥ 1 year | 5 | 0.73 (0.52-1.05) | 0% | 0.09 | 0.07 | 5 | 0.72 (0.42-1.25) | 0% | 0.25 | 0.79 | 7 | 0.88 (0.69-1.13) | 54% | 0.32 | 0.94 | 6 | 0.59 (0.35-1.00) | 16% | 0.05 | 0.02 | 4 | 0.18 (0.04-0.77) | 0% | 0.02 | 0.09 |
| < 1 year | 5 | 1.08 (0.86-1.34) | 0% | 0.51 |  | 3 | 0.65 (0.39-1.10) | 0% | 0.92 |  | 4 | 0.90 (0.59-1.35) | 0% | 0.52 |  | 3 | 1.39 (0.88-2.19) | 0% | 0.16 |  | 2 | 1.11 (0.25-4.96) | 0% | 0.89 |  |
| Sample size |  |  |  |  |  |  |  |  |  |  |  |  |  |  |  |  |  |  |  |  |  |  |  |  |  |
| ≥ 100 | 5 | 1.02 (0.84-1.23) | 0% | 0.85 | 0.09 | 4 | 0.72 (0.48-1.08) | 0% | 0.11 | 0.49 | 3 | 0.75 (0.52-1.07) | 60% | 0.12 | 0.26 | 4 | 1.04 (0.72-1.50) | 61% | 0.83 | 0.21 | 3 | 0.27 (0.07-1.05) | 16% | 0.06 | 0.83 |
| < 100 | 5 | 0.46 (0.19-1.12) | 0% | 0.09 |  | 4 | 0.47 (0.15-1.46) | 0% | 0.19 |  | 8 | 0.97 (0.74-1.26) | 0% | 0.81 |  | 5 | 0.54 (0.21-1.40) | 0% | 0.21 |  | 3 | 0.48 (0.11-2.02) | 0% | 0.31 |  |
| Concomitant antiplatelet medication |  |  |  |  |  |  |  |  |  |  |  |  |  |  |  |  |  |  |  |  |  |  |  |  |  |
| Complete<sup>#</sup> | 2 | 0.51 (0.09-3.04) | 0% | 0.46 | 0.48 | 2 | 0.59 (0.10-3.60) | 0% | 0.57 | 0.88 | 2 | 0.76 (0.50-1.17) | 76% | 0.22 | 0.43 | 2 | 0.37 (0.17-0.81) | 0% | 0.01 | 0.006 | 2 | 0.14 (0.02-1.00) | 0% | 0.05 | 0.2 |
| Incomplete | 8 | 0.97 (0.81-1.17) | 0% | 0.77 |  | 6 | 0.69 (0.47-1.01) | 0% | 0.06 |  | 9 | 0.93 (0.73-1.19) | 9% | 0.56 |  | 7 | 1.25 (0.85-1.85) | 0% | 0.26 |  | 4 | 0.61 (0.19-1.97) | 0% | 0.41 |  |
| Primary causes of LVT |  |  |  |  |  |  |  |  |  |  |  |  |  |  |  |  |  |  |  |  |  |  |  |  |  |
| MI | 2 | 0.34 (0.10-1.25) | 0% | 0.11 | 0.09 | 1 | 0.49 (0.05-4.53) | - | 0.63 | 0.76 | 2 | 0.57 (0.38-0.84) | 0% | 0.005 | 0.006 | 2 | 0.22 (0.03-1.67) | 0% | 0.14 | 0.14 | 2 | 0.14 (0.02-1.03) | 0% | 0.05 | 0.23 |
| Mixed | 8 | 1.00 (0.83-1.21) | 0% | 0.96 |  | 7 | 0.69 (0.47-1.01) | 0% | 0.96 |  | 9 | 1.09 (0.85-1.41) | 0% | 0.47 |  | 7 | 1.02 (0.72-1.44) | 26% | 0.91 |  | 4 | 0.57 (0.18-1.85) | 0% | 0.35 |  |

<sup>#</sup> Complete medication indicates all patients of a study receive at least one antiplatelet agent along with anticoagulants; incomplete medication indicates studies with some of patients receiving no antiplatelet agents or not reporting
3 data of antiplatelet medications. CI = confidence interval, LVT = left ventricular thrombus, N = number of studies in the subgroup, NOACs = novel oral anticoagulants, MI = myocardial infarction, P<sub>int</sub> = P for interaction, RR = risk ratio, SSE = stroke
4 or systemic embolism, VKAs = vitamin K antagonists.

**Figure 2.**
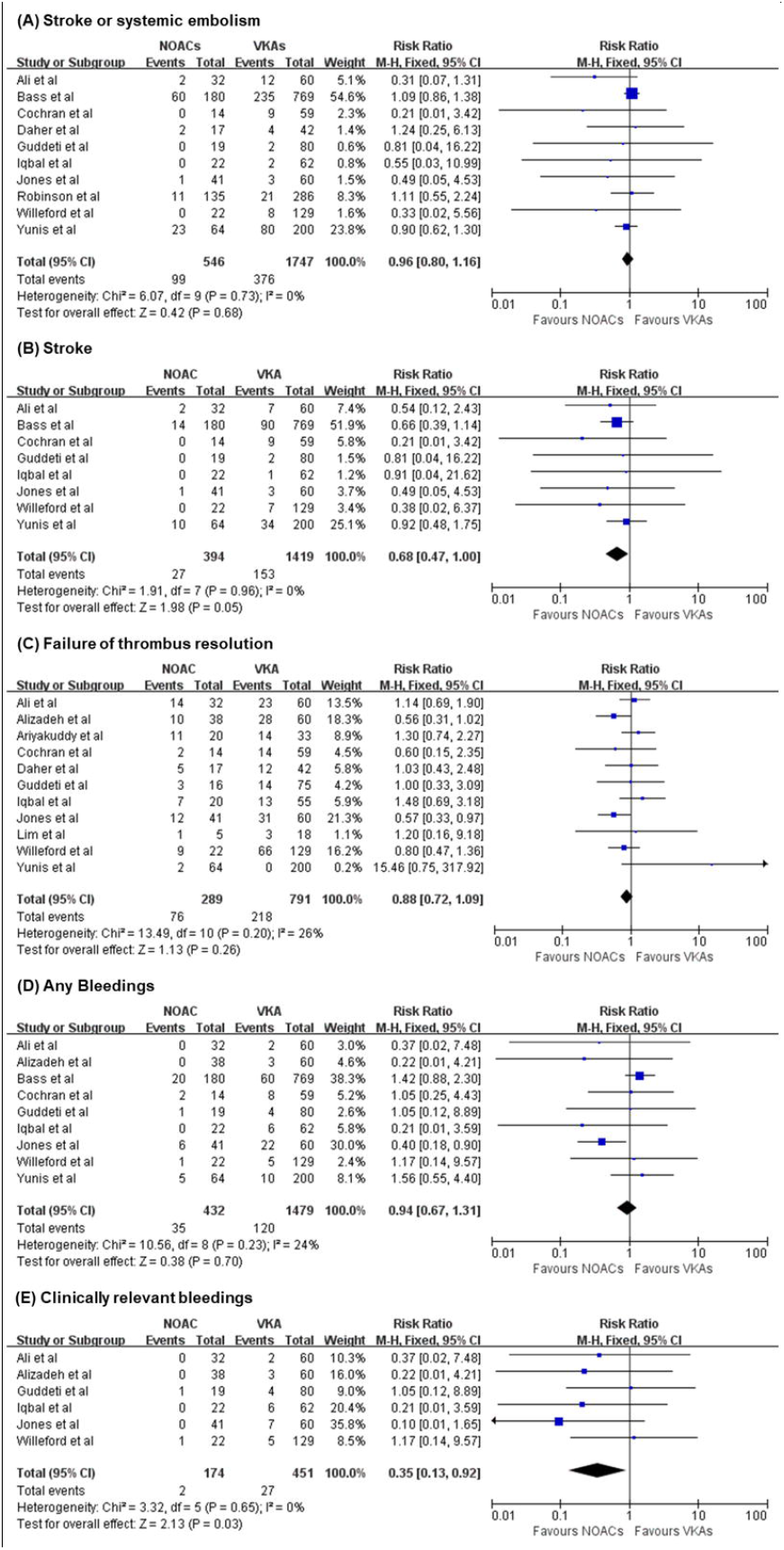
Forest plots showing pooled risk ratio for users of novel oral anticoagulants (NOACs) versus vitamin K antagonists (VKA) regarding outcomes of stroke or systemic embolism (A), stroke (B), failure of thrombus resolution (C), any bleedings (D), and clinically relevant bleedings (E).

### Thrombus resolution

Eleven studies investigated the outcome of failure in thrombus resolution ^5-9,16-19,21,22,24,25^, and the resolution rate was similar in two groups (RR: 0.88, 95% CI: 0.72-1.09, P = 0.20, **Figure 2C**), with an low statistical heterogeneity (I^2^ = 26%, P = 0.20) and no significant publication bias based on the funnel plot (**Supplementary figure 1C**) and statistical tests (Begg’s test, P = 0.484; Egger’s test, P = 0.191). Age did not show substantial impacts on thrombus resolution according to meta-regression (RR: 1.08, 95% CI: 0.95-1.22, P = 0.215, **Supplementary figure 3C**). Efficacy for thrombus resolution was consistent despite follow-up duration (P _interaction_ = 0.94), sample size (P _interaction_ = 0.26) and anti-platelet medication (P _interaction_ = 0.43) in subgroup analysis (**Table 2**). However, significant interactions were observed for primary causes for LVT (myocardial infarction [MI], RR: 0.57, 95% CI: 0.38-0.84, I^2^ = 0%, P = 0.005; mixed etiologies, RR: 1.09, 95% CI: 0.85-1.41, I^2^ =0%, P = 0.47; P _interaction_ = 0.006)

### Bleeding events

Nine studies reported bleeding events^6,9,16,19-22,24,25^, the risk of any bleedings was similar for NOACs and VKAs users (RR: 0.94, 95% CI: 0.67-1.31, P = 0.70; I^2^ = 24%, **Figure 2D**), without significant bias for publication as shown in the funnel plot (**Supplementary figure 1D)**. Meta-regression showed age did not differentiate the safety of NOACs or VKAs (RR: 1.03, 95% CI: 0.81-1.31, P = 0.764, **Supplementary figure 3D**). Subgroup analysis (**Table 2**) showed bleeding risk was similar despite sample size (P _interaction_ = 0.21) and etiologies (P _interaction_ = 0.14). However, significant interactions were observed for follow-up duration (P _interaction_ = 0.02) and antiplatelet medications (P _interaction_ = 0.006).

In six studies reporting clinically relevant bleedings ^6,9,16,22,24,25^, NOACs users showed more favorable outcomes (RR: 0.35, 95% CI: 0.13-0.92, P = 0.03; I^2^ = 0, **Figure 2E**), and no significant publication bias was detected by the funnel plot (**Supplementary figure 1E**). Age did not affect the difference in clinically relevant bleeding risk between NOACs and VKAs (RR: 0.83, 95% CI:0.41-1.69, P = 0.516, **Supplementary figure 3E**). Subgroup analysis showed no significant interactions with follow-up duration (P _interaction_ = 0.09), sample size (P _interaction_ = 0.83), antiplatelet strategies (P _interaction_ = 0.2) and etiologies (P _interaction_ = 0.23) for risk for clinically relevant bleedings of using NOACs or VKAs (**Table 2**).

## Discussions

The major findings of this systematic review and meta-analysis are as follow: (1) only observational studies have been conducted regarding anticoagulation treatment for LVT, and most of them are retrospective, (2) NOACs users showed similar risk of SSE, failure of LVT resolution, but lower risk of strokes compared with VKAs users, and (3) NOACs users acquired similar risk of any bleedings but lower risk of clinically relevant bleedings as compared with VKAs users.

### Efficacy of NOACs and VKAs

According to the current guidelines, warfarin is still recommended as the first-line treatment for LVT, although with no evidence from RCTs ^1,3,4^. However, NOACs is gaining interest for LVT treatments, as it acquires more consistent anticoagulant effects while reducing bleeding risks ^1,26^. Growing numbers of cases and clinical studies also suggest a satisfactory outcome for LVT patients using NOACs ^6-9^ . Although the current analysis suggested no difference between NOACs and VKAs for their efficacy in SSE prevention, as reported by other recently published meta-analysis ^21,27^, it was definitely of great clinical importance as it showed a 32% risk reduction for strokes alone in NOACs users. Subgroup analysis also showed a highly homogenous reduction in the risk of stroke across various confounders. A recently published pooled-analysis by Zhou et al shows no difference in stroke (odds ratio: 0.79, 95% CI: 0.50-1.23) for NOACs and VKAs users ^27^, but the current studies included three newly published research in this year ^21,24,25^, which enlarged the sample size and provided greater power to test the difference between two medications. Although the interpretation was challenging, one of the main reasons could be the fluctuation of the INR. Ali et al report that 71% of stroke patients receiving warfarin have suboptimal control INR ^25^. In the study by Jones et al ^9^, nearly half of warfarin users could not sustain an ideal INR for over 65% of time during the coagulation treatment, of whom 75% were below the target value, while all the thromboembolic events occurred in those patients with sub-optimally controlled INR. Therefore, physicians should consider initiating NOACs treatment to provide long-term consistent anticoagulation for LVT patients, especially when there are difficulties in monitoring or sustaining INR in an ideal range. Additionally, it should be noticed that there are some discrepancies among included studies, like the study by Robinson et al reports a substantial increase in risk of SSE in NOACs (HR: 2.67, 95% CI: 1.31-5.57) users in their original article, which is quite the opposite to other included studies ^23^. However, up to 15% of their patients switched anticoagulants during the follow-up, making it hard to estimate the true risk difference between NOACs and warfarin. In two recent studies on the same topic, impacts from this issue are less discussed ^21,27^. In the current analysis taking an intention-to-treat approach, the inclusion and exclusion of this study did not bring much variation to the pooled effect and heterogeneity, which affirmed the neutral results in pooled analysis. Taken together, NOACs did not increase the risk of SSE for LVT patients, and effectively reduced the risk of stroke as compared with VKAs, possibly due to its more constant anticoagulation effects. Moreover, the current study showed thrombus resolution rate was similar for NOACs and VKAs users, which was consistent with previous reports ^21,27^. Interestingly, MI patients receiving NOACs showed significantly higher rates of thrombus resolution compared to those using VKAs. These discrepancies could be due to the increased thrombotic burden after MI. For MI patients, LVT is dynamically formed within a few days after the initial cardiac damage ^28,29^. Moreover, the exposure of subendothelial contents due to myocardial necrosis intensified the prothrombotic states, which could sustain as long as six months ^1,30^. Therefore, it is reasonable to start on effective and constant anticoagulation as early as possible to limit the progress of thrombus formation. However, effects of VKAs are expected to peak after 72 to 96 hours after initial dosage before the existing clotting factors are depleted ^31,32^, while suboptimal control of INR is frequently reported in users of warfarin ^9,25^. Such disadvantages could have constituted a less pro-resolutive environment for MI patients challenged by actually greater thrombotic burden compared to LVT patients with other etiologies. In sum, NOACs and VKAs generally offered similar efficacy of thrombus resolution, while NOACs could be a more suitable choice for choice for patients with MI.

### Safety of NOACs and VKAs

The current analysis showed that NOACs did not reduce the risk of any bleedings of LVT patients, but significantly lowered the risk of clinically relevant bleedings as compared with VKAs. These findings were generally consistent with previous studies regarding patients with atrial fibrillation and heart failure ^10,11^. Notably, NOACs showed homogenous reduction across various subgroups for clinically relevant bleedings, which could be very important for LVT patients. Although the life-threatening bleeding rate is quite low for various anticoagulant agents ^5-9^, there is still increasing need for greater reduction of major bleedings, as oral anticoagulants are frequently prescribed to LVT patients on top of antiplatelet agents due to complex etiologies and background diseases ^1,3,4^, which inevitably and substantially increases the actual bleeding risk. For any bleedings, despite the neutral pooled effects, NOACs still achieved lower bleeding rates for longer follow-up and patients taking antiplatelet medications, which is beneficial for improving life quality and adherence of patients in need of long-term anticoagulation or antiplatelet treatments ^33,34^. Taken together, NOACs still possessed a better safety profile than VKAs, as it effectively reduced clinically important bleeding events, and showed a potential to reduce overall bleeding rates in patients taking longer and more intensified antithrombotic treatment.

## Limitations

The general limitations of this meta-analysis are as follow. Firstly, the included studies were all observational. Although baseline characteristics were generally consistent in NOACs and VKAs group in most of the studies, confounding effects could be present due to lack of adjustment from individual data. Secondly, the NOACs regime and dosage were different in each study. Thirdly, bleeding outcomes were not reported according to standard definitions (like BARC classification). Future well-designed RCTs could be safely initiated to assess efficacy of NOACs and VKAs for LVT patients based on the evidence from current analysis ^35^.

## Conclusions

For patients with LVT, the efficacy and safety profile were similar between NOACs and VKAs. However, NOACs effectively lowered the risk of stroke and clinically relevant bleedings as compared with VKAs. Therefore, NOACs could be safely initiated in patients with LVT as primary oral anticoagulants.

## Data Availability

Data for this analysis could be acquired through the original publications.

## Author contributions

RC and HY contributed to the conception or design of the work. RC, JZ, CL, PZ, JL, XZ, YW, LS and HZ contributed to the acquisition, analysis, or interpretation of data for the work. RC, JZ and LS drafted the manuscript. RC, HZ, HY critically revised the manuscript. All authors gave final approval and agree to be accountable for all aspects of work ensuring integrity and accuracy

## Conflicts of Interest

The authors declare that they have no conflicts of interest.

### Acknowledgments

We sincerely thanked Dr Raviteja R. Guddeti (Creighton University School of Medicine) for helping us to accomplish the current analysis by providing necessary outcomes data, and Dr Weida Liu (Medical Research & Biometrics Center, Fuwai Hospital) for his professional advice in statistical analysis.

