## Supplementary tables and figures for "Novel Oral Anticoagulants versus Vitamin K Antagonists for Patients with Left Ventricular Thrombus: A Systematic Review and Meta-Analysis"

**Supplementary Material**

**Supplementary table 1.** Scoring details for included studies by Newcastle-Ottawa quality assessment scale for cohort studies.

| Study | Selection | | | |  | Comparability |  | Outcome | | |  | Total |
| --- | --- | --- | --- | --- | --- | --- | --- | --- | --- | --- | --- | --- |
|  | (1) | (2) | (3) | (4) |  | (1) |  | (1) | (2) | (3) |  |  |
| Ali et al | ※ | ※ | ※ | ※ |  | No adjustment |  | ※ | ※ | ※ |  | 7 |
| Alizadeh et al | ※ | ※ | ※ | ND |  | ※※ |  | ND | ※ | ND |  | 4 |
| Ariyakuddy et al | ※ | ※ | ※ | ND |  | No adjustment |  | ※ | ※ | ND |  | 5 |
| Bass et al | ※ | ※ | ※ | ※ |  | No adjustment |  | ND | ※ | ND |  | 5 |
| Cochran et al | ※ | ※ | ※ | ND |  | ※※ |  | ※ | ※ | ※ |  | 8 |
| Dasher et al | ※ | ※ | ※ | ※ |  | No adjustment |  | ※ | ※ | ND |  | 5 |
| Guddeti et al | ※ | ※ | ※ | ND |  | No adjustment |  | ND | ※ | ※ |  | 5 |
| Iqbal et al | ※ | ※ | ※ | ND |  | No adjustment |  | ND | ※ | ※ |  | 5 |
| Jones et al | ※ | ※ | ※ | ND |  | ※※ |  | ※ | ※ | ※ |  | 8 |
| Lim et al | ※ | ※ | ※ | ND |  | No adjustment |  | ND | ※ | ※ |  | 5 |
| Robinson et al | ※ | ※ | ※ | ※ |  | ※※ |  | ND | ※ | ※ |  | 7 |
| Willeford et al | ※ | ※ | ※ | ※ |  | ※※ |  | ※ | ※ | ※ |  | 9 |
| Yunis et al | ※ | ※ | ※ | ND |  | ※※ |  | ND | ※ | ※ |  | 7 |
| Average | - | - | - | - |  | - |  | - | - | - |  | 6.2 |

ND = no description.

**Supplementary table 2.** Solution of unclear outcome data of stroke or systemic embolism in the study by Robinson et al.

| Patients allocation  (NOAC vs Warfarin) | Pooled risk ratio (95% CI) | I^2^ | P-value |
| --- | --- | --- | --- |
| 134 vs 287 | 0.96 [0.80, 1.16] | 0 | 0.68 |
| 135 vs 286 | 0.96 [0.80, 1.16] | 0 | 0.68 |
| 136 vs 285 | 0.96 [0.80, 1.16] | 0 | 0.67 |
| 137 vs 284 | 0.96 [0.80, 1.16] | 0 | 0.66 |
| 138 vs 283 | 0.96 [0.80, 1.15] | 0 | 0.66 |
| 139 vs 282 | 0.96 [0.80, 1.15] | 0 | 0.65 |
| Current study excluded | 0.95 [0.78, 1.15] | 0 | 0.59 |

In the study by Robinson et al, a total of 64 (15.2%) out of 421 patients underwent treatment switches between novel oral anticoagulants (NOACs) and warfarin. Therefore, we took the intention-to-treat approach for this study in the pooled analysis. Although authors have provided data regarding the switching, the original anticoagulants could not be determined in seven patients who switching twice between NOACs and warfarin. We have tried contacting the authors to obtain these data, but no response so far. Therefore, we tried different allocations for these seven patients to the group of NOACs and warfarin (i.e. 1&6, 2&5, 3&4), and the pooled risk ratio were very similar (see table below), which suggested limited impacts of different combinations. The result sustained after excluding the study. Therefore, we allocated these seven patients proportionally to two groups, namely two patients to NOACs and five patients to warfarin, and the pooled risk ratio was 0.96 (95% CI: 0.80-1.16, P=0.68, I^2^=0%). Thrombus resolution and bleeding events were only categorized according to the anticoagulants being used at the incidence of the index events instead of original anticoagulants, and therefore the pooled analysis for these two outcomes excluded the data from the current study.

**Supplementary figure 1.** The funnel plots for outcomes of stroke or systemic embolism (A), stroke (B), failure of thrombus resolution (C), any bleedings (D), and clinically relevant bleedings (E).

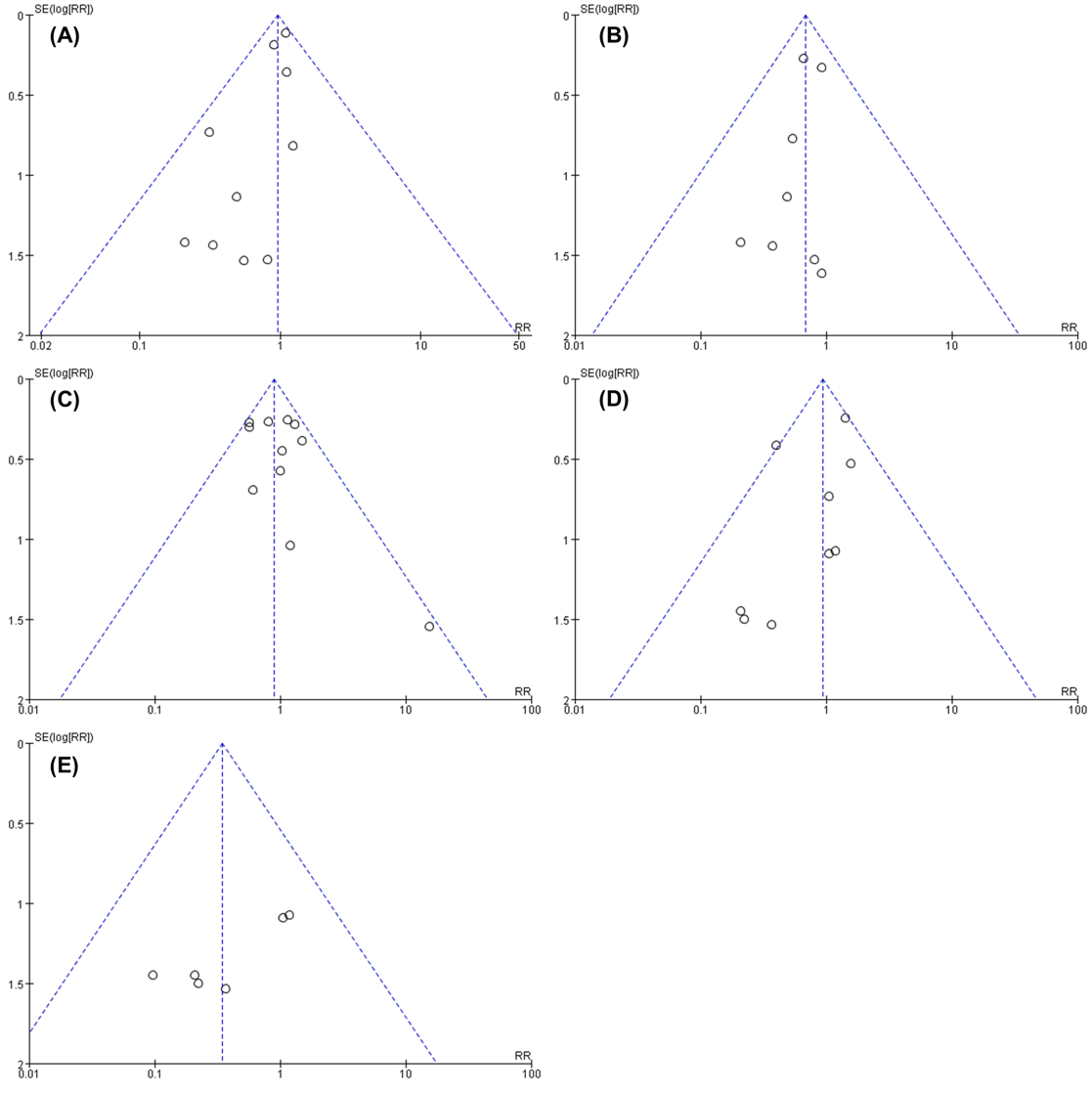

**Supplementary figure 2.** The funnel plot after imputation by trim-and-fill methods for the outcome of stroke or systemic embolism. No extra studies were imputed for the current analysis.

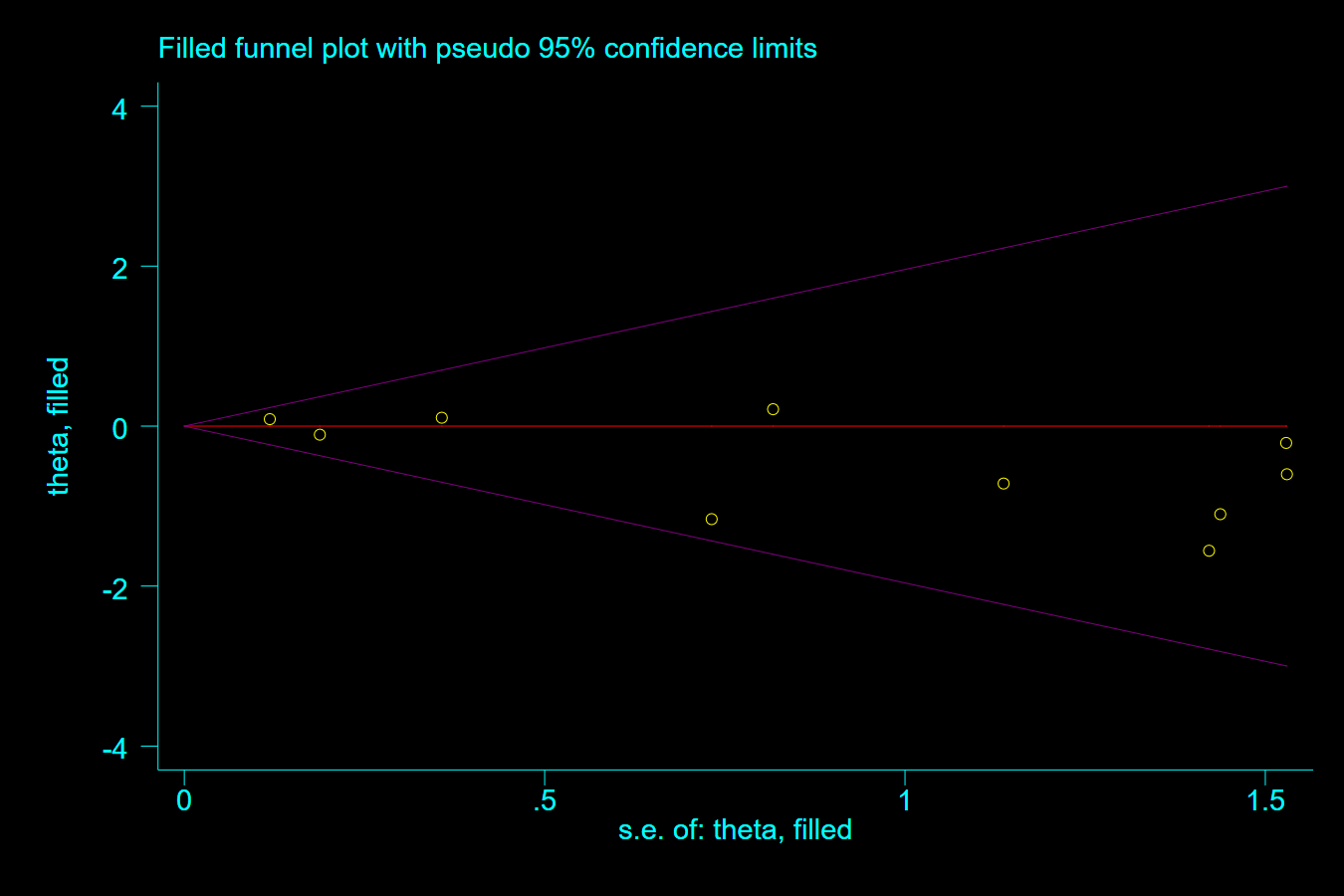

**Supplementary figure 3**. Meta-regression of risk ratio (RR) against average or median age according to included studies for outcomes of stroke or systemic embolism (A), stroke (B), failure of thrombus resolution (C), any bleedings (D), and clinically relevant bleedings (E).

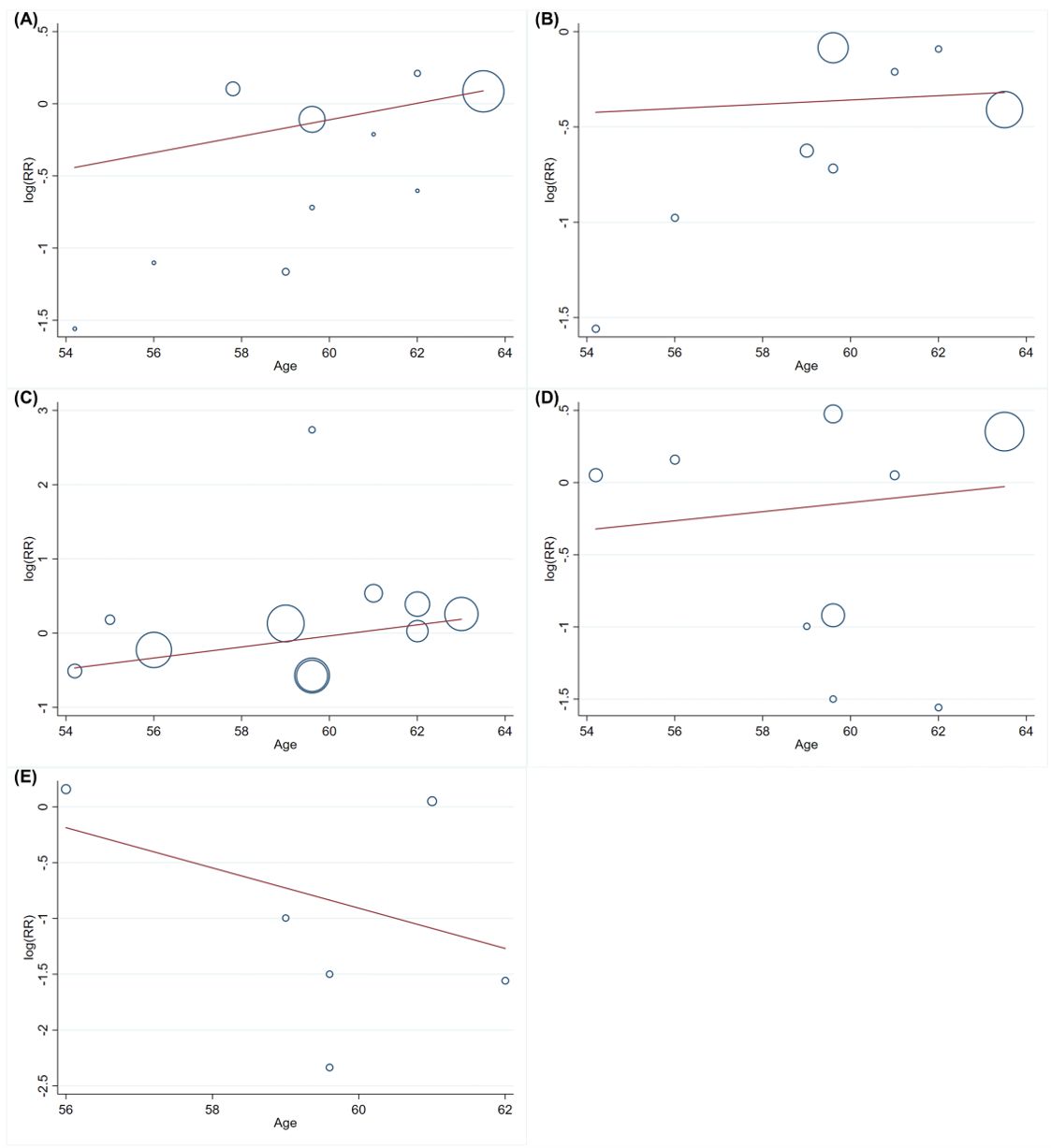
